# Do intramural career development programs provide an advantage to rehabilitation scientists applying for merit-review scientific funding? A retrospective cohort study

**DOI:** 10.1101/2025.08.27.25334557

**Authors:** John J. Fraser, Eric J Schwinder, Erin D Spaniol

## Abstract

**Introduction:** The Veterans Health Administration (VHA) offers intramural Career Development Awards (CDAs). The purpose of this research was to investigate if completing a VHA CDA influenced the funding success rate for subsequent VHA Merit Review award applications in rehabilitation science.

**Methods:** A retrospective cohort study of all applicants who submitted a VHA Merit Review award application to the Rehabilitation Research, Development, and Translation (RRDT) Broad Portfolio from fiscal year 2020 to 2025 was performed. The factors of experience as a prior CDA recipient and the number of application submissions on the success of funding was evaluated in early investigators and mid-career/senior scientists.

**Results:** Early-career investigators with prior VHA CDAs (n=14) submitted 32 new/revised applications for VHA Merit Review award funding. While early investigators with prior VHA CDAs had a numerically higher percentage of funding across the three application submission cycles (34.4% vs 25.0%), this factor was not statistically significant. Mid-career and senior investigators with prior VHA CDAs (n=78) submitted 180 new/revised applications and demonstrated an overall 30.0% funding success across the three-application submission cycle. Mid-career and senior investigators with prior VHA CDAs had a statistically significant increased odds of funding across the three-application submission cycle (30.0% vs 26.4%; OR_unadj_:1.62; OR_adj_: 1.54).

**Discussion:** While investigators with prior VHA CDAs did not have a significant advantage when applying for VHA Merit Review awards from ORD’s RRDT Portfolio during the early-career phase, there was a statistically significant benefit observed in mid-career and senior scientists on the initial application submission or overall.

**Conclusion:** Based on the potential benefits of the mentored research experience within the VA Medical Centers observed in the current study and in preceding studies, early-career scientists interested in working with Veterans are encouraged to apply for these awards following completion of the terminal degree or post-doctoral fellowship.

## INTRODUCTION

As the number of individuals that live longer and with chronic diseases and injuries increase, there is a need for investment in rehabilitation health services to improve the function and health-related quality of life of the population.^1,2^ This includes Veterans that have served in the armed forces, a population whose prevalence of service-connected disabilities have doubled in the past decade.^3^ In response to the growth of adults who are older or have chronic medical conditions, the World Health Organization detailed in the “Rehabilitation 2030: a call for action” position statement the need to scale up rehabilitation capabilities, to include the number of rehabilitation scientists.^2^ One program that serves as a generator of rehabilitation research capabilities are Career Development Award (CDA) programs. The Veterans Health Administration (VHA) offers CDAs (https://www.research.va.gov/funding/CDP.cfm) as an intramural funding mechanism through the Office of Research and Development (ORD). These awards provide early-career scientists (including clinicians and non-clinician) protected time for continued didactic and experiential training in a structured mentored research program. These programs allow the continued development of research knowledge, skills, abilities needed to be a competitive independent investigator that extends beyond academic and clinical doctoral and post-doctoral training, which are often times limited in time and scope.^4^ In the scoping review conducted by Li and colleagues,^5^ the authors detailed the importance of protected time and individualized mentorship to acquire relevant expertise and academic competence needed for a successful research career. VHA offers CDAs at two levels. The CDA-1 is designed for junior-level investigators who recently concluded academic training and are beginning their postdoctoral research careers.

It consists primarily of two years of salary support and is structured on close collaboration with a primary mentor who holds concurrent VHA research funding. It is expected that by the second year of the CDA-1, program participants will apply for the next level of VHA CDA funding, the CDA-2. The CDA-2 builds upon a research foundation (gained during the CDA-1, via one or more fellowships, or other postdoctoral research training). The CDA-2 is targeted towards early-career scientists who have demonstrated their potential for independent research. The award includes salary support and project funds. Participants work with a team of mentors who support the recipient’s scientific progress, productivity, and training, and are expected to apply for independent research funding (from VHA or other sources) in the final two years of their funding period. This analysis examines the effectiveness of these programs by analyzing the rates at which CDA-2 participants go on to secure VHA independent research funding (the VHA Merit Review Award). All funding applications submitted to VHA ORD are evaluated for scientific merit by a federal advisory committee comprised of government and non-government subject matter experts, followed by a programmatic review. For both CDAs and Merit Review mechanisms, applicants may revise and resubmit unsuccessful applications up to two times (for a total of three submissions per application).

While the importance of rehabilitation scientists participating in career development programs have been articulated in commentary^4,6^ or study of perceived value,^7^ to the authors’ knowledge there has not been an empirical evaluation of these programs on subsequent funding success in rehabilitation scientists. Therefore, the purpose of this research was to investigate whether obtaining and completing a VHA CDA influenced the success rate of rehabilitation scientists applying for VHA Merit Review award funding in the ORD Rehabilitation Research, Development, and Translation (RRDT) Broad Portfolio (https://www.research.va.gov/isrm/rrdt/, formerly known as the Rehabilitation Research and Development Service prior to 2025). Due to the selection of only the most qualified applicants, the structured didactics and experiential learning opportunities, mentored research experience, development of scientific expertise in areas with the greatest need, and the time to develop relationships, research networks, and esoteric corporate knowledge, we posit that early-career scientists with VHA CDA experience will have increased success in their subsequent applications for VHA research funding, requiring fewer submission attempts compared to early investigators who did not participate in a VHA CDA. While it is plausible that the effect on success rates would persist beyond the early-career stage, we speculate that these benefits will washout when the study population is broadened to include all investigators who applied for funding.

## METHODS

A retrospective cohort study of all applicants who submitted a VHA Merit Review award application to the ORD RRDT Broad Portfolio from fiscal year 2020 to 2025 was performed (Winter funding cycle only for 2025). The factors of experience as a prior CDA recipient (referenced to investigators who did not benefit from a VA CDA) and the number of application submissions (first and final resubmissions referenced to the initial submission) on the success of funding was evaluated in early investigators (≤10-years research experience), mid-career/senior scientists (>10-years research experience), and in all merit review award applicants. This study was determined to be non-human subjects research by the VA Central Institutional Review Board. The Strengthening the Reporting of Observational Studies in Epidemiology (STROBE) guidelines were used to guide reporting.^8^

The count of distinct investigators applying for a Merit Review award, initial and revised application submissions, and funded awards in ORD’s RRDT Broad Portfolio was derived from the VHA’s Research Analysis and Forecasting Tool (RAFT) database (VA Office of Research and Development, Washington, DC), an internal platform used to manage research funds and track expenditures. Status as an early-career investigator was derived from Electronic Research Administration (eRA) Commons (National Institute of Health [NIH], Bethesda, MD), an online interface used for managing research grant applications and awards.^9^

Funding percentage, unadjusted odds ratios (OR), and chi-squared statistics were calculated using Microsoft® Excel for Mac (version 16.98, Microsoft Corp., Redmond, WA) and a custom epidemiological spreadsheet.^10^ Multivariable logistic regression models were employed to assess the adjusted OR of prior VHA CDA experience, the interaction of career stage and prior VHA CDA, the number of Merit Review application submissions, and career stage on award funding success using R (version 4.5, The R Foundation for Statistical Computing, Vienna, Austria). The level of significance was *p* < 0.05 for all analyses. OR point estimates were considered statistically significant if CIs did not cross the 1.00 threshold. Convergence of p-values, effect sizes, and 95% confidence intervals were considered when evaluating significant findings.

## RESULTS

Early-career investigators with prior VHA CDAs (n=14) submitted 32 new/revised applications for VHA Merit Review award funding and demonstrated an overall 34.4% funding success across the three application submission cycles (n=Initial: 14.3%; First resubmission: 20.0%; Final resubmission: 87.5%) (**Table 1**). Early-career investigators without prior VHA CDAs (n=22) submitted 40 new/revised applications and demonstrated an overall 25.0% funding success across the three-application submission cycle (Initial: 0.0%; First resubmission: 46.2%; Final resubmission: 80.0%). While early-career investigators with prior VHA CDAs had a numerically higher percentage of funding in the three submission cycles, this factor was not statistically significant in the unadjusted (OR_unadj_:1.75) or adjusted analyses (OR_adj_: 0.99) (**Figure 1**). There was a statistically significant increased odds of success in the first (OR_adj_: 9.07) and final resubmissions (OR_adj_: 93.75) in all early-career investigators (**Table 4**).

**Figure 1.**
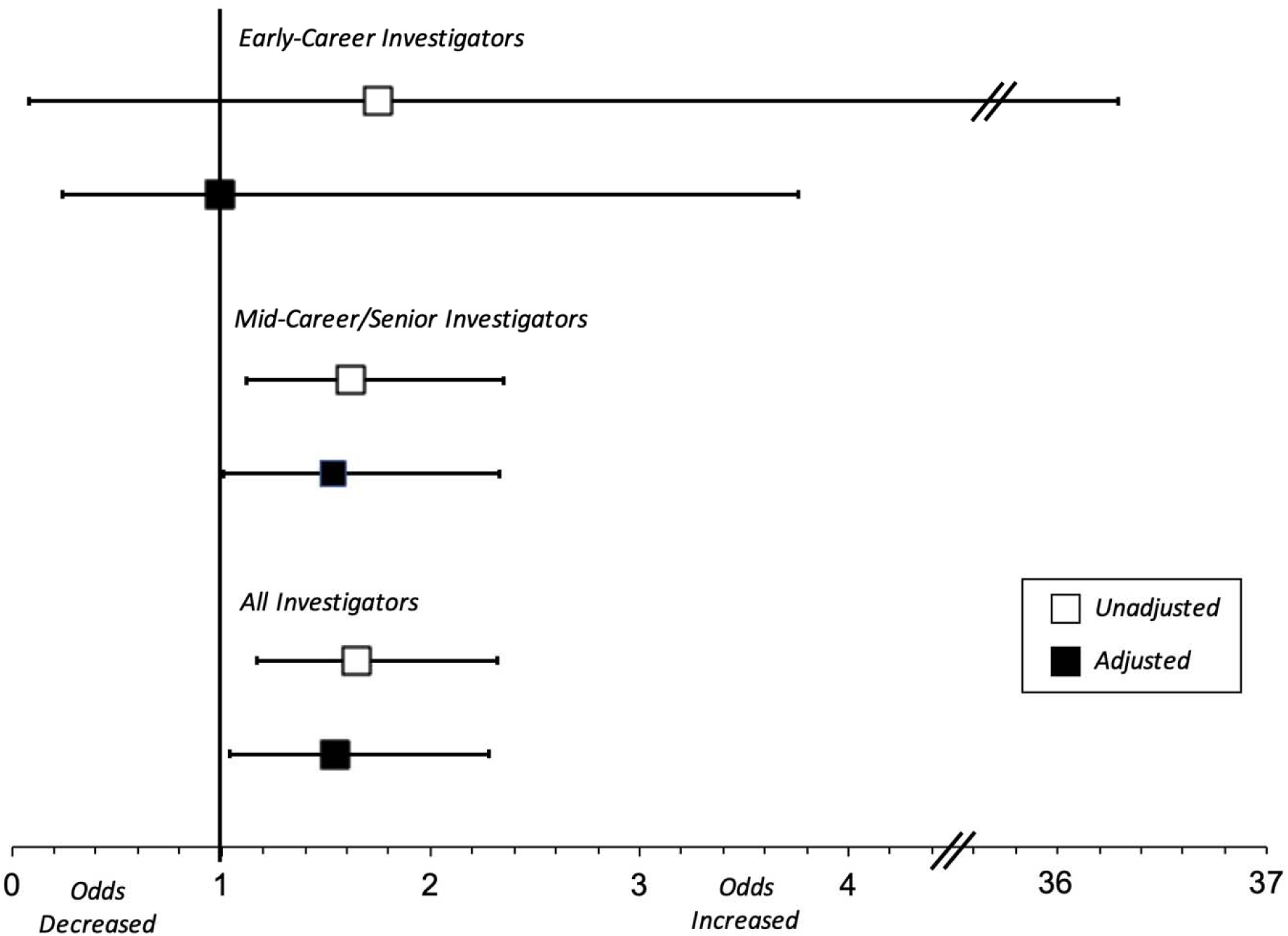
Unadjusted and adjusted odds of merit review funding success in rehabilitation investigators with a prior Veterans Health Administration Career Development Award referenced to those without.

**Table 1.** Counts, probability, and unadjusted odds ratios of awards in the prior Career Development Award (CDA) and non-CDA groups, stratified by submission, in early-career scientists applying for Veterans Affairs merit awards, 2020-2025.

|  |  | Merit Award Submission |  |  | Overall |
| --- | --- | --- | --- | --- | --- |
|  |  | Initial | First Resubmission | Final Resubmission |  |
| Prior-CDA Group | Awards (n) | 2 | 2 | 7 | 11 |
|  | Applications (n) | 14 | 10 | 8 | 32 |
|  | Funding % | 14.3% | 20.0% | 87.5% | 34.4% |
| Non-CDA Group | Awards (n) | 0 | 6 | 4 | 10 |
|  | Applications (n) | 22 | 13 | 5 | 40 |
|  | Funding % | 0.0% | 46.2% | 80.0% | 25.0% |
| Odds Ratios (non-CDA reference) |  | 36.7 | 0.29 | 8.75 | 1.75 |
| 95% CI |  | 0.06-21844.57 | 0.04-1.94 | 0.74-103.82 | 0.08-36.29 |
| p-value |  | † | † | † | .72 |
| * 0.1 substituted for 0 count. † Chi-square statistic inappropriate for counts <5. CI, confidence interval; NS, non-significant. |  |  |  |  |  |

Mid-career and senior investigators with prior VHA CDAs (n=78) submitted 180 new/revised applications and demonstrated an overall 30.0% funding success across the three-application submission cycle (Initial: 11.5%; First resubmission: 35.9%; Final resubmission: 57.9%) (**Table 2**). Mid-career and senior investigators without prior VHA CDAs (n=318) submitted 640 new/revised applications and demonstrated an overall 20.9% funding success across the three-application submission cycle (Initial: 5.0%; First resubmission: 26.4%; Final resubmission: 56.4%). Mid-career and senior investigators with prior VHA CDAs had a statistically significant increased odds of funding in the initial submission of new applications (OR_unadj_: 2.46) and across the three-application submission cycle (OR_unadj_:1.62; OR_adj_: 1.54) (**Figure 1**). There was a statistically significant increased odds of success in the first (OR_adj_: 5.90) and final resubmissions (OR_adj_: 19.30) in mid-career and senior investigators (**Table 4**).

**Table 2.** Counts, probability, and unadjusted odds ratios of awards in the prior Career Development Award (CDA) and non-CDA groups, stratified by submission, in mid-career and senior scientists applying for Veterans Affairs merit awards, 2020-2025.

|  |  | Merit Award Submission |  |  | Overall |
| --- | --- | --- | --- | --- | --- |
|  |  | Initial | First Resubmission | Final Resubmission |  |
| Prior-CDA Group | Awards (n) | 9 | 23 | 22 | 54 |
|  | Applications (n) | 78 | 64 | 38 | 180 |
|  | Funding % | 11.5% | 35.9% | 57.9% | 30.0% |
| Non-CDA Group | Awards (n) | 16 | 56 | 62 | 134 |
|  | Applications (n) | 318 | 212 | 110 | 640 |
|  | Funding % | 5.0% | 26.4% | 56.4% | 20.9% |
| Odds Ratios (non-CDA reference) |  | **2.46** | 1.56 | 1.06 | **1.62** |
| 95% CI |  | **1.04-5.80** | 0.86-2.83 | 0.50-2.24 | **1.12-2.35** |
| p-value |  | **.03** | NS | NS | **.01** |

Bolded values indicate statistical significance. CI, confidence interval; NS, non-significant.

In the pooled evaluation of all investigators, agnostic of career stage, investigators with prior VHA CDAs (n=92) submitted 212 new/revised applications and demonstrated an overall 30.7% funding success across the three-application submission cycle (Initial: 12.0%; First resubmission: 33.8%; Final resubmission: 63.0%) (**Table 3**). Investigators without prior VHA CDAs (n=340) submitted 680 new/revised applications and demonstrated an overall 21.2% funding success across the three-application submission cycle (Initial: 4.7%; First resubmission: 27.6%; Final resubmission: 57.4%). Investigators with prior VHA CDAs had a statistically significant increased odds of funding in the initial submission of new applications (OR_unadj_: 2.75) and across the three-application submission cycle (OR_unadj_:1.65; OR_adj_: 1.54) (**Figure 1**). There was a statistically significant increased odds of success in the first (OR_adj_: 6.11) and final resubmissions (OR_adj_: 21.31) (**Table 4**). There were no other significant findings.

**Table 3.** Counts, probability, and unadjusted odds ratios of award in the prior Career Development Award (CDA) and non-CDA groups, stratified by submission, in all scientists applying for Veterans Affairs merit awards, 2020-2025.

|  |  | Merit Award Submission |  |  | Overall |
| --- | --- | --- | --- | --- | --- |
|  |  | Initial | First Resubmission | Final Resubmission |  |
| Prior-CDA Group | Awards (n) | 11 | 25 | 29 | 65 |
|  | Applications (n) | 92 | 74 | 46 | 212 |
|  | Funding % | 12.0% | 33.8% | 63.0% | 30.7% |
| Non-CDA Group | Awards (n) | 16 | 62 | 66 | 144 |
|  | Applications (n) | 340 | 225 | 115 | 680 |
|  | Funding % | 4.7% | 27.6% | 57.4% | 21.2% |
| Odds Ratios (non-CDA reference) |  | **2.75** | 1.34 | 1.27 | **1.65** |
| 95% CI |  | **1.23-6.15** | 0.76-2.36 | 0.63-2.56 | **1.17-2.32** |
| p-value |  | **.01** | NS | NS | **.004** |

Bolded values indicate statistical significance. CI, confidence interval; NS, non-significant.

**Table 4.**
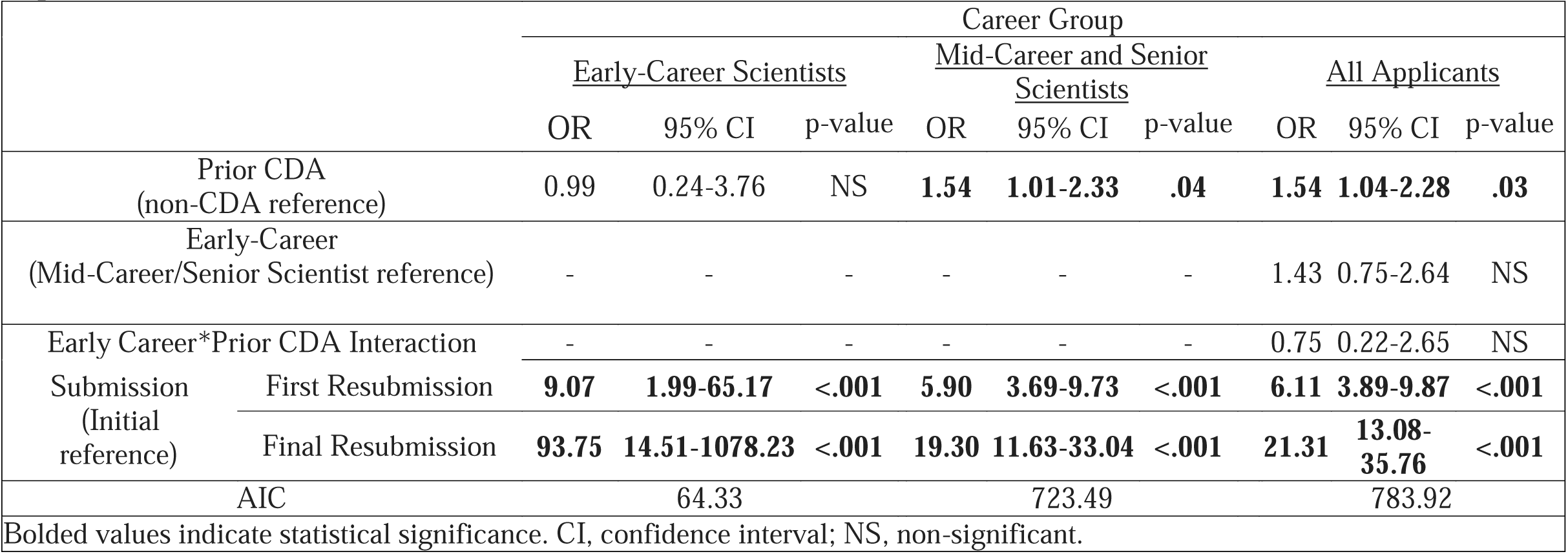
Adjusted odds ratios of awards in the prior Career Development Award (CDA) and non-CDA groups, stratified by career stage.

## DISCUSSION

The primary finding of this study was that while investigators with prior VHA CDAs did not have a significant advantage over other applicants without this experience when applying for VHA Merit Review awards from ORD’s RRDT Portfolio during the early-career phase, there was a statistically significant benefit observed in mid-career and senior scientists on the initial application submission or overall. This is the first study to the authors’ knowledge to have investigated the effects of the VHA CDA program on VHA Merit Review funding success rates in rehabilitation scientists.

VHA CDA experience appears to influence career trajectory in intramural rehabilitation scientists with 10 or more years of experience. There are some plausible reasons for this observation. Due the competitive nature of VHA CDA awards and the selection of candidates with the greatest potential and expressed commitment to the VA, selection bias may explain the observed advantage. However, while the adage of prior success as a predictor of future performance may apply in many contexts, it appears to be not the case in merit review grant funding.^11^ In a retrospective evaluation of NIH R01 grant funding over a 15-year epoch, Prassad and colleagues^11^ found that prior funding was not predictive for future grant funding, with funding success in both high- and low-performers observed to have regressed toward the mean.

Another plausible explanation for our observations may be related to the number of years an intramural investigator has served in the VHA research enterprise. Due to the time required to apply (and resubmit), meet administration requirements, receive funding, and execute on a VHA CDA, investigators with these awards have the advantage years of experience within the organization before competing for a merit review award. In the phenomenon known as the “The Matthew Effect,” scientists may have competitive advantage due to the social relationships, associations, and notoriety developed in the organization and the broader field of study, especially among early-career scientists.^12,13^ It is plausible that performance on the VHA CDA also provided additional time in service to the VHA needed to develop salient scientific lines of study that are of value to Veterans and the broader organization. These suppositions warrant future investigation.

Based on the potential benefits of the mentored research experience observed in the current study and in preceding studies,^14–16^ early-career scientists interested in working with Veterans are encouraged to apply for these awards following completion of the terminal degree or post-doctoral fellowship. We also acknowledge that early-career scientists need the support of a strong mentorship team and the facility in where they serve. Established career scientists and organizational leaders are encouraged to serve as mentors and support career development programs in the recruitment and training of early-investigators, who serve as the life-blood and the future generation of innovators in the VA. There are limitations to this study. We observed a relatively small sample size in the early-investigator group during the 5-year cohort, especially among applicants with a prior CDA.

In eRA, the 10-year period indicating early-career status starts from the date of their terminal research degree or the end of post-graduate clinical training.^9^ Applicants with prior CDAs may start applying for merit review applications in the latter few years of their early-career stage, which likely curtailed the number of applicants in this category. While there is a risk that a type II error was committed (indicating a false negative), comfort may be taken when considering that any difference would be of relatively small magnitude (9.4% greater overall funding percentage of prior CDAs over their non-CDA counterparts). The delimitations in this study required that investigators in the prior-CDA group needed have this training award specifically funded by VA ORD. It is likely that some intramural investigators included in the non-prior CDA group may have benefitted from a training award from another institution, such as the NIH K-award mechanism. While both VA and non-VA CDA mechanisms provide mentored research experiences, it is plausible that investigators that trained specifically under a VA CDA benefited from experiences esoteric to the VA enterprise and additional knowledge, skills, abilities, and esoteric knowledge unique to the care of Veterans. Future research is needed to validate this supposition.

## CONCLUSION

Mid-career and senior rehabilitation scientists with a prior intramural CDA had significantly greater odds of obtaining merit-review research funding on their initial application and across the three-cycles compared to their counterparts who did not have this developmental experience. It is unclear if this benefit was applicable in early-career scientists with a CDA due to limitations of this study. The investment of a mentored experiential research program during the early-career stage appears to have careerlong benefits in rehabilitation scientists applying for intramural funding in the VA.

## Supporting information

Executive Summary

## Data Availability

All data produced in the present study are available upon reasonable request to the authors.

## Acknowledgements

Charmaine O’Brien, MS for her assistance in data extraction and administrative support.

