## Supplementary material for "Do intramural career development programs provide an advantage to rehabilitation scientists applying for merit-review scientific funding? A retrospective cohort study": Executive Summary

Fraser JJ, Schwinder EJ, Spaniol ED. Do intramural career development programs provide an advantage to rehabilitation scientists applying for merit-review scientific funding? A retrospective cohort study. Office of Research & Development, US Department of Veterans Affairs. Unpublished report. 2025.

**BLUF:** Mid-career and senior rehabilitation scientists with a prior intramural VHA Career Development Award had significantly greater odds of obtaining merit-review research funding on their initial application and across the three-cycles compared to their counterparts who did not have this developmental experience.

**Background:** The Veterans Health Administration (VHA) offers Career Development Awards (CDAs) as an intramural funding mechanism through its Office of Research and Development (ORD). These awards provide early-career scientists (including clinicians and non-clinician) protected time for continued didactic and experiential training in a structured mentored research program. While the importance of rehabilitation scientists participating in career development programs have been articulated in commentary^1,2^ or study of perceived value,^3^ there has not been an empirical evaluation of these programs on subsequent funding success in rehabilitation scientists. Therefore, the purpose of this research was to investigate whether successfully obtaining and completing a VHA CDA influenced the funding success rate for subsequent VHA Merit Review award applications in rehabilitation science.

**Methods:** A retrospective cohort study of all applicants, consisting of both early-career ( ≤10-years experience) and mid-career/senior (>10-years experience) scientists, who submitted a VHA Merit Review award application to ORD’s Rehabilitation Research, Development, and Translation (RRDT) Broad Portfolio (formerly known as the Rehabilitation Research and Development Service) from fiscal year 2020 to 2025 was performed. Crude and adjusted odds ratios (OR) were used to evaluate merit-review funding success in applicants with a prior CDA referenced to those without this experience.

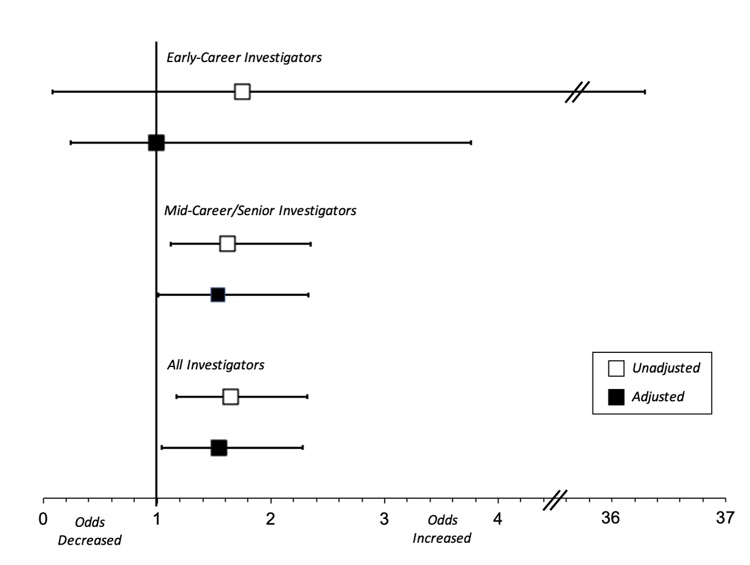
**Results**: Investigators with prior VHA CDAs (n=92) submitted 212 new/revised applications and demonstrated an overall 30.7% funding success across the three-application submission cycle (Initial: 12.0%; First resubmission: 33.8%; Final resubmission: 63.0%) Investigators without prior VHA CDAs (n=340) submitted 680 new/revised applications and demonstrated an overall 21.2% funding success across the three-application submission cycle (Initial: 4.7%; First resubmission: 27.6%; Final resubmission: 57.4%). Investigators with prior VHA CDAs had a statistically significant increased odds of funding in the initial submission of new applications (OR_unadj_: 2.75) and across the three-application submission cycle (OR_unadj_:1.65; OR_adj_: 1.54) (**Figure**).

**Discussion**: This is the first study to the authors’ knowledge to have investigated the effects of the VHA CDA program on VHA Merit Review funding success rates in rehabilitation scientists. Based on the potential benefits of the mentored research experience observed in the current study and in preceding studies,^4,5^ early career scientists interested in working with Veterans are encouraged to apply for these awards following completion of the terminal degree or post-doctoral fellowship. We also acknowledge that early-career scientists need the support of a strong mentorship team and the facility in where they serve. Established career scientists and organizational leaders are encouraged to serve as mentors and support career development programs in the recruitment and training of early-investigators, who serve as the life-blood and the future generation of innovators in the VA. Policies that incentivize VA Medical Centers to develop and foster CDA programs allow for talent growth and force-shaping based on the evolving needs of Veterans, and VHA capabilities as a research and healthcare enterprise.

**Disclaimer:** The authors are employees of the U.S. Government and this work was prepared as part of their official duties. Title 17, U.S.C. §105 provides that copyright protection under this title is not available for any work of the U.S. Government. Title 17, U.S.C. §101 defines a U.S. Government work as work prepared by a military service member or employee of the U.S. Government as part of that person’s official duties. The views expressed are those of the authors and do not necessarily reflect the official policy or position of the Department of Veterans Affairs, nor the U.S. Government.
